# Quantifying Cause-Effect Relations Between Walking Speed, Propulsive Force, and Metabolic Cost

**DOI:** 10.1101/2021.10.18.21265129

**Authors:** Richard E. Pimentel, Jordan N. Feldman, Michael D. Lewek, Jason R. Franz

## Abstract

Walking speed is a useful surrogate for health status across the population. Walking speed appears to be governed in part by propulsive force (F_P_) generated during push-off and simultaneously optimized to minimize metabolic cost. However, no study to our knowledge has established empirical cause-effect relations between F_P_, walking speed, and metabolic cost, even in young adults. To overcome the potential linkage between these factors, we used a self-paced treadmill controller and real-time biofeedback to independently prescribe walking speed or F_P_ across a range of condition intensities. Walking with larger and smaller F_P_ led to instinctively faster and slower walking speeds, respectively, with about 80% of variance explained between those outcomes. We also found that comparable changes in either F_P_ or walking speed elicited predictable and relatively uniform changes in metabolic cost, each explaining about ∼53% of the variance in net metabolic power and ∼15% of the variance in cost of transport, respectively. These findings build confidence that interventions designed to increase F_P_ will translate to improved walking speed. Repeating this protocol in other populations may identify additional cause-effect relations that could inform the time course of gait decline due to age and disease.

## Introduction

Walking speed serves as a simple surrogate for human health status. For example, faster walking speeds associate with a host of positive health factors, including increased muscle strength, better cognitive function, greater independence, and reduced healthcare costs.^1–7^ By understanding the mechanistic pathways that contribute to slower walking speeds we may identify avenues to maintain and restore independence and pedestrianism for safe and effective recreation, transport, and health in our population.

We often attribute the selection of walking speed to the minimization of metabolic cost. The cost of transport (CoT, *i*.*e*., net metabolic cost per unit distance traveled) during walking is U-shaped, with increasing costs as walking speed deviates from preferred. This suggests that our movement biomechanics and underlying muscle actions are tuned to minimize metabolic cost at our preferred speeds. Unfortunately, compared to young adults or unimpaired controls, numerous walking studies in older adults or people with gait limitations document higher CoT^8–12^ and slower preferred walking speeds.^11,12^ A variety of factors likely explain the higher CoT in these individuals, including systemic factors (e.g., reduced cardiopulmonary function), local muscle and tendon factors (e.g., reduced muscle metabolic efficiency, lower tendon stiffness), and altered neural control or gait biomechanics (e.g., wider steps, increased co-activation, redistributing mechanical work to more proximal leg joints/muscles). However, the often-simultaneous presentation of slower speeds and higher CoT challenges our ability to fully understand the time course of gait decline due to aging or gait pathology.

Biomechanically, walking speed is understood to be regulated by the magnitude of the peak anterior component of the ground reaction force^13^ - namely, the peak propulsive force (F_P_). We generate F_P_ from the trailing limb during push-off, which acts to accelerate and redirect the body’s center of mass forward and upward, thereby regulating walking speed. Humans typically generate a vigorous F_P_ via ankle plantarflexion using a combination of well-timed calf muscle contraction, elastic energy returned from the Achilles tendon, and effective limb orientation for mechanical advantage.^14–18^ Because the ankle plantarflexor muscles and tendons account for about 60% of the work performed in typical gait,^14,19^ it is no anomaly that plantarflexor pathologies affect both walking speed and walking economy.^8,11,20^ This suggests that F_P_, walking speed, and metabolic cost are inextricably linked, posing a longstanding scientific challenge with significant potential for improved clinical countermeasures.

Before scientists and clinicians can design and implement strategies to improve walking speed and lower metabolic cost in older adults or in individuals with gait pathology, we need to better understand exactly how F_P_ impacts walking speed in the context of walking metabolic cost. To our knowledge, no study has established empirical cause-effect relations between F_P_, walking speed, and walking metabolic cost, even in unimpaired young adults. Thus, our purpose was to: (1) investigate how F_P_ governs the selection of walking speed, and (2) quantify how the selection of F_P_ or walking speed impacts walking economy. Exploring these relations may build confidence that restoring F_P_ will lead to improvements in walking speed. Additionally, our results may be useful when designing interventions or devices that seek to improve walking ability and economy.

## Methods

### Participants

We recruited a convenience sample of twenty young adults who provided informed consent prior to any activities in this study. The University of North Carolina at Chapel Hill institutional review board approved all research procedures. All participants were free of current lower extremity injuries, neuromuscular complications, and walking assistive devices that might prevent protocol completion. On average, participants were 24.7 ± 5.2 years old *(mean ± standard deviation)*, stood at a height of 1.77 ± 0.11 m, had a mass of 75.6 ± 13.7 kg, and thus had a BMI of 24.0 ± 3.4 kg/m^2^.

### Self-Pace Mode and Targeted F_P_ Biofeedback

Understanding our experimental design depends on understanding the self-pace treadmill controller and targeted biofeedback we implemented. Thus, we first describe our techniques and follow with our protocol. This study leveraged a self-paced treadmill controller adapted from Hedrick et al. (2021).^17^ Trials using self-pacing always started at the participant’s preferred overground walking speed (see below), following which they could instantaneously increase or decrease the treadmill speed at will by moving towards the front or back of the treadmill, respectively (Fig. 1). We used participants’ average center of pressure position during each double support phase to determine their relative anterior-posterior location on our force-sensing, dual belt treadmill (Bertec Corp., Columbus, Ohio, USA). When the average center of pressure position moved beyond the 20 cm “dead zone” centered on the treadmill midline, walking speed changed linearly with the distance from center:

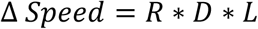

with R indicating the relative sign of the speed change (−1 when posterior to dead zone, and +1 when anterior to dead zone), D defining the average center of pressure distance from the treadmill center, and L linearly scaling the speed change, set to 0.1 in this study based on pilot testing.

**Figure 1:**
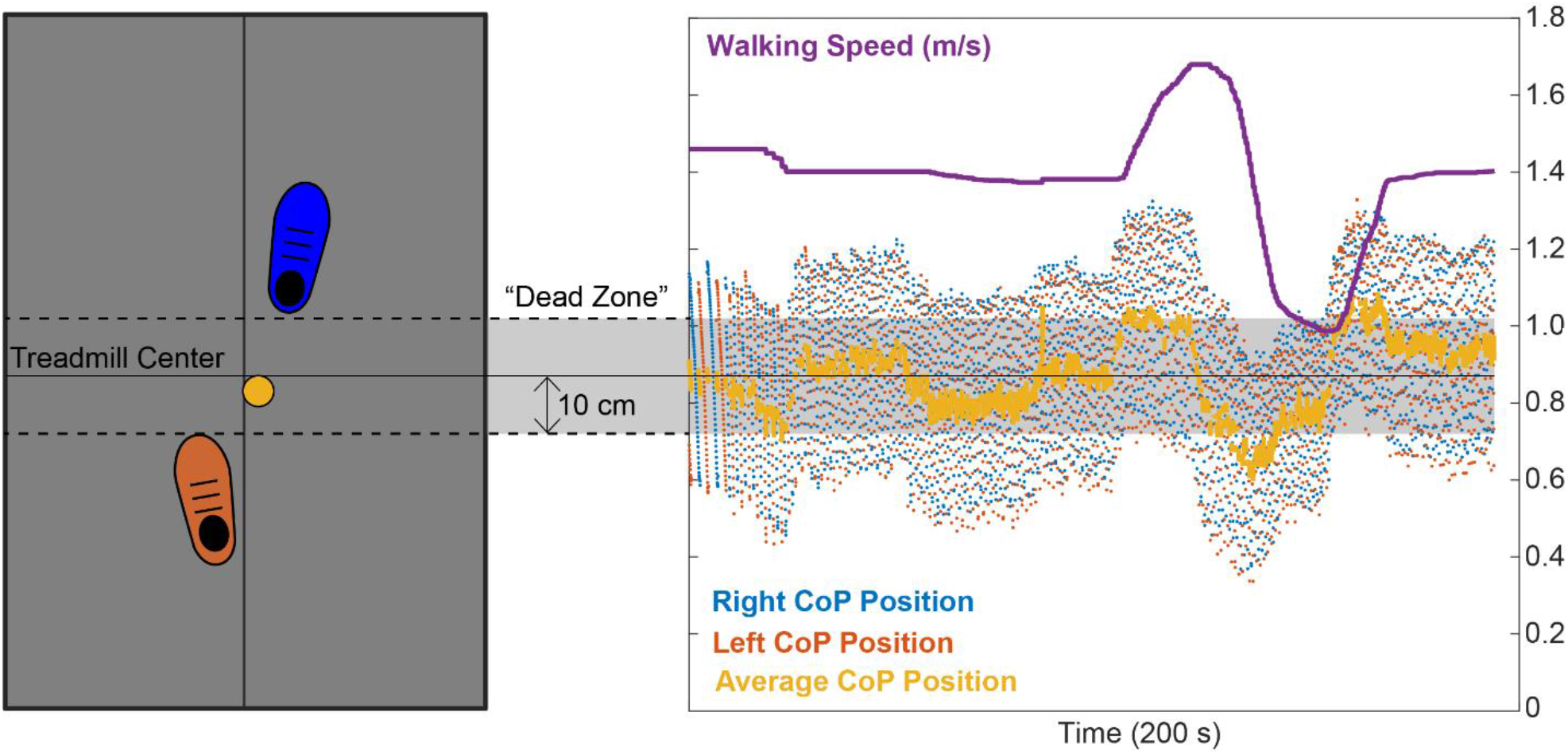
In self-pace mode, participants started walking on a split-belt treadmill at their preferred overground walking speed. Using a Matlab script, we recorded participants’ instantaneous bilateral centers of pressure (CoP) from each belt in real time and averaged the sides to estimate their relative fore/aft position on the treadmill (yellow dot & line). When the participant stayed centered on the treadmill (i.e., average CoP location during double support within the 20 cm “dead zone”), treadmill speed remained constant. When the participant’s average CoP location during double support moved anterior or posterior to the dead zone, the treadmill speed increased or decreased linearly with the distance from center, respectively.

In some trials, we also used real-time visual biofeedback to display the average peak F_P_ from the previous two steps on a screen in front of the participant with a target line representing the prescribed F_P_ according to our study protocol (see below). We instructed participants to “match their push off force to the target”. The biofeedback line turned green when participants’ instantaneous F_P_ for that step was within 5% of the target value (in newtons), but was otherwise red. For additional encouragement, we provided a counter displaying the number of consecutive steps on target. We provide the treadmill controller scripts, written in Matlab (Mathworks, Natick, MA, USA) and using the software development kit between Cortex (Motion Analysis Corportaion, Rohnert Park, CA, USA) and Matlab at github.com/peruvianox/FpBiofeedbackSelfPace.

### Experimental Protocol

We determined participants’ preferred walking speed via the average speed from four 30-m passes in a hallway where we instructed them to “walk normally, as if you were walking down a sidewalk”. Participants completed a 3-minute warm-up walk at their preferred walking speed followed by a 3-minute walk to become familiar with the self-pacing treadmill mode and targeted F_P_ biofeedback. During the self-pace and targeted biofeedback warm-up, we ensured all participants could increase and decrease their F_P_ on command and could regulate their walking speed at will using the self-pace mode.

Figure 2 summarizes our experimental protocol. Participants walked at a fixed speed (speed clamp) for five 5-minute trials at their preferred speed (Norm) and at ±10% and ±20% of Norm in randomized order. We extracted the average peak F_P_ from each speed clamp trial to use as targets for the ensuing biofeedback trials. Participants then completed a randomized series of 5-minute walking trials with biofeedback to target their average peak F_P_ from each of the speed clamp trials. These targeted biofeedback trials used the self-paced treadmill controller, thus prescribing a target F_P_ while allowing walking speed to vary (aptly-named: F_P_ clamp). Participants rested in a seated position for at least 1 minute between trials, and were allowed to take longer rests ad libitum (average time between trials: 167±105 s).

**Figure 2:**
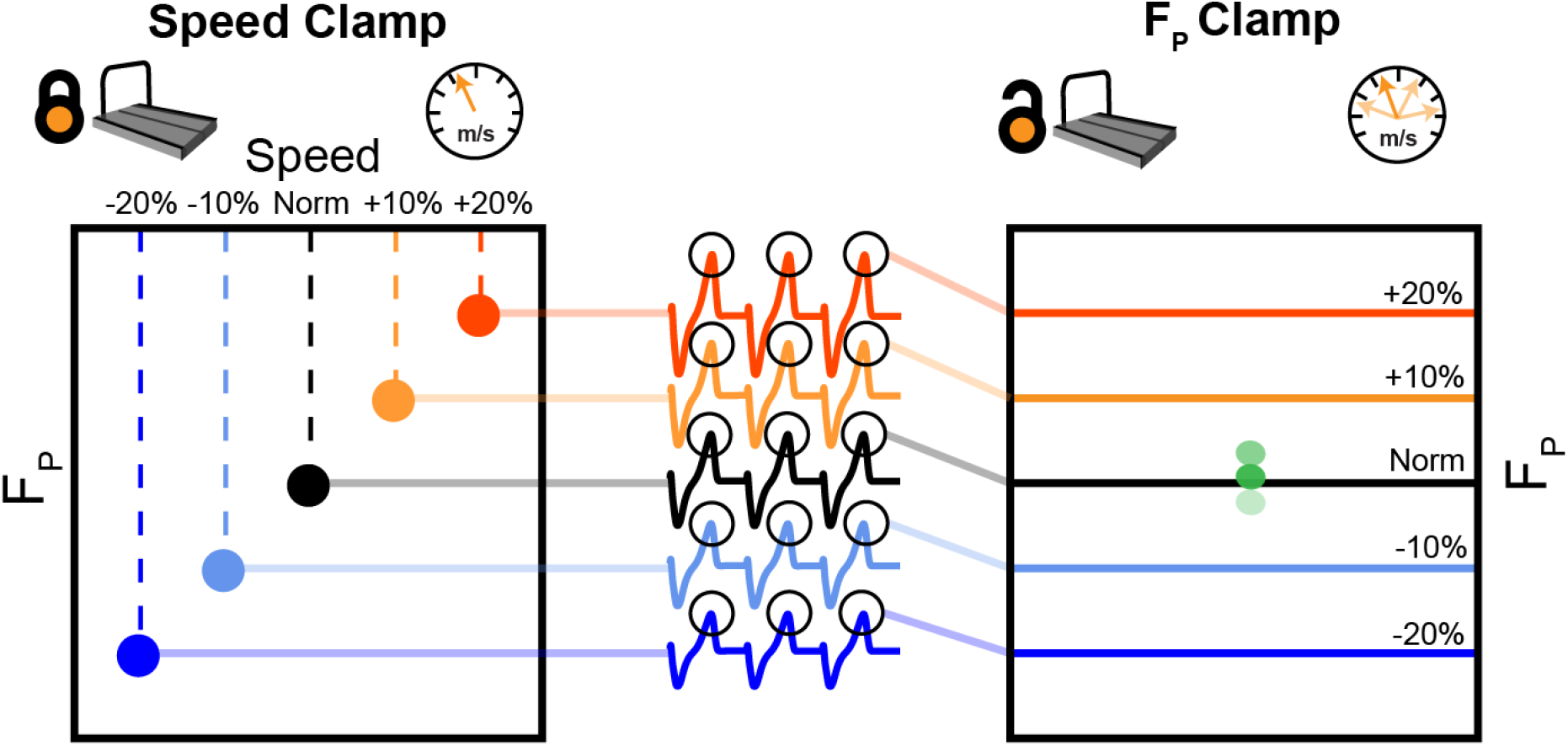
Participants first walked at their typical, overground walking speed (Norm) as well as ±10% and ±20% of Norm. During these 5-minute, fixed-speed trials (speed clamp), we measured and averaged F_P_ from each step over the duration of the trial. During another set of five 5-minute trials, we used targeted biofeedback and the self-paced treadmill mode to prescribe F_P_ by asking participants to target each of their average F_P_ values from the speed clamp while allowing participants to naturally adjust their walking speed (F_P_ clamp).

We recorded treadmill speed and ground reaction forces from the real-time treadmill controller script. The real-time interface between Cortex (Motion Analysis Corporation, Rohnert Park, CA, USA) and Matlab (Mathworks, Natick, MA, USA) received data packets every 0.050 ± 0.002 s, with 10 embedded analog force samples at 1000 Hz included in each packet. We analyzed average treadmill speed and bilateral average peak F_P_ over the final two minutes of each trial for statistical analysis, allowing participants to explore and stabilize their walking patterns for the first 3 minutes of each trial.

### Metabolic Measurements

In a baseline standing trial and all walking trials, we sampled exhaled oxygen and carbon dioxide on a breath-by-breath basis using a COSMED K5 portable metabolic system (COSMED, Rome, Italy) via indirect calorimetry. To estimate standing and walking net metabolic power, we respectively averaged expired air measurements over the final two minutes of the 5-minute standing collection and each 5-minute walking trial. Standard regression equations estimated whole-body metabolic power from rates of oxygen consumption and carbon dioxide production.^21^ We subtracted standing metabolic power from walking metabolic power to calculate net metabolic power, and lastly, normalized by body mass.

### Data Analysis and Statistics

We squared Pearson correlations to quantify the relations between F_P_, walking speed, and metabolic outcomes (i.e., net metabolic power and CoT). We considered coefficient of determination strengths using the following classification (>0.8=very strong, 0.6-0.8=strong, 0.4-0.6=moderate, 0.2-0.4=weak, <0.2=very weak). We used a two-way repeated measures analysis of variance (ANOVA) to identify main effects of and interactions between clamp type (speed vs. F_P_) and condition intensity (Norm, ±10%, ±20%) on walking speed, F_P_, net metabolic power, and CoT. When we found a significant main effect or interaction, we used Tukey’s post-hoc tests to identify pairwise differences factors and conditions. We provide effect sizes for comparison of all significant statistical outcomes – with η_p_^2^ for ANOVA main effects and Cohen’s d for post-hoc comparisons. We performed all statistical processing in python using the [Pingouin](https://pingouin-stats.org/) package. For transparency and open science initiative, we provide our data files and analysis code at github.com/peruvianox/SpeedFpClamp.

## Results

### Participants and Experimental Performance

On average, participants walked at a typical overground speed of 1.41±0.09 m/s and propelled themselves forward during treadmill walking with a typical F_P_ of 22.0±2.3 % body weight (*mean ± standard deviation*). Figure 3 shows (A) F_P_ during speed clamp trials, (B) F_P_ clamp trial biofeedback targeting performance, and (C) subsequent changes in walking speed during F_P_ clamp trials. In summary, these panels demonstrate that participants modified their F_P_ according to prescribed targets with accompanying changes in walking speed.

**Figure 3:**
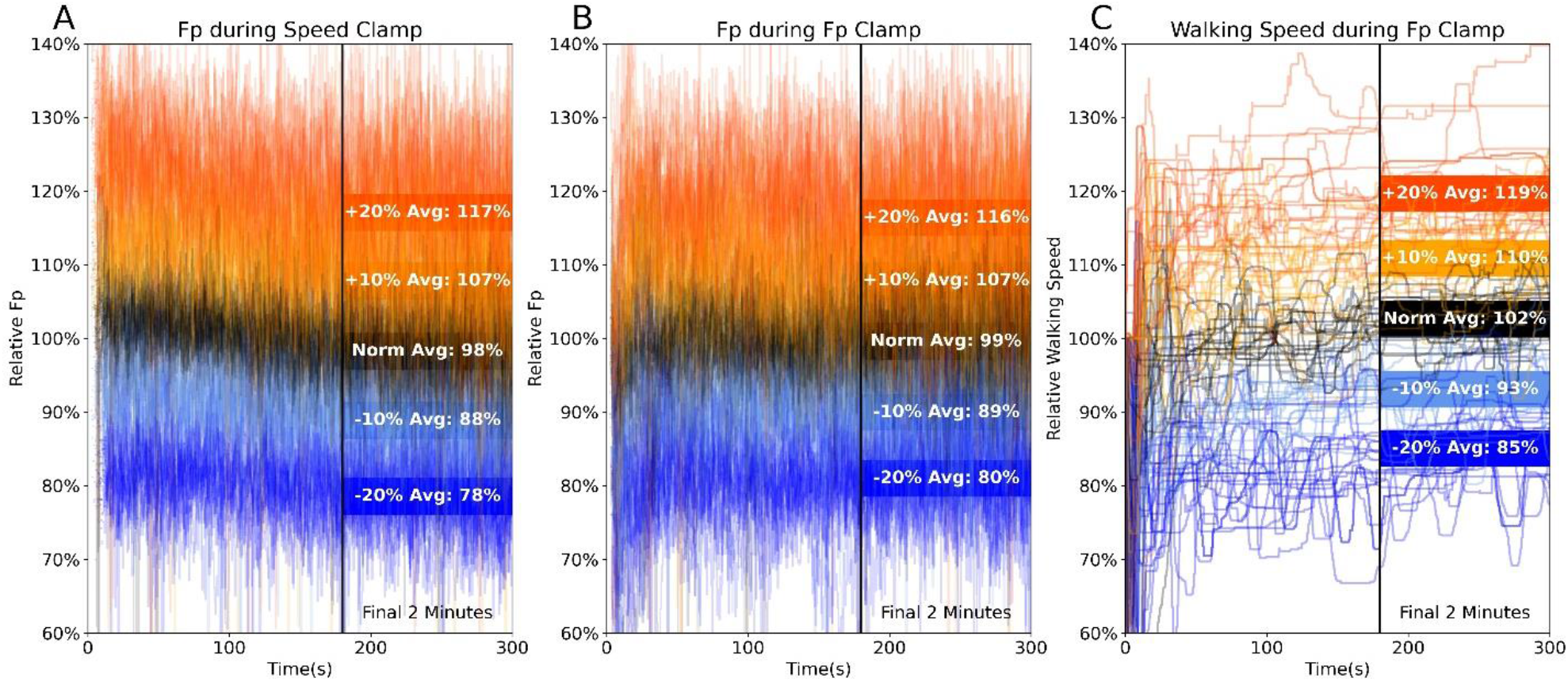
For every step during all trials and across all subjects, we show relative F_P_ for the speed clamp (A), F_P_ clamp trial biofeedback targeting performance (B), and instantaneous self-paced walking speed relative to usual walking (C). Overall, participant’s average F_P_ and walking speed over the final two minutes generally agreed well with prescribed changes in condition intensity. Early variation in relative walking speed stabilized after the first ∼90s.

### Correlations

Across both clamp types, walking speed very strongly correlated with F_P_, (average R^2^=0.80, p<0.001, Fig. 4A), moderately correlated with net metabolic power (R^2^=0.54, p<0.001, Fig. 4B), and very weakly correlated with CoT (R^2^=0.13, p<0.001, Fig. 4C). Similarly, across both clamp types, F_P_ moderately correlated with net metabolic power (R^2^=0.52, p<0.001, Fig. 4D) and very weakly correlated with CoT (R^2^=0.17, p<0.001, Fig. 4E).

**Figure 4:**
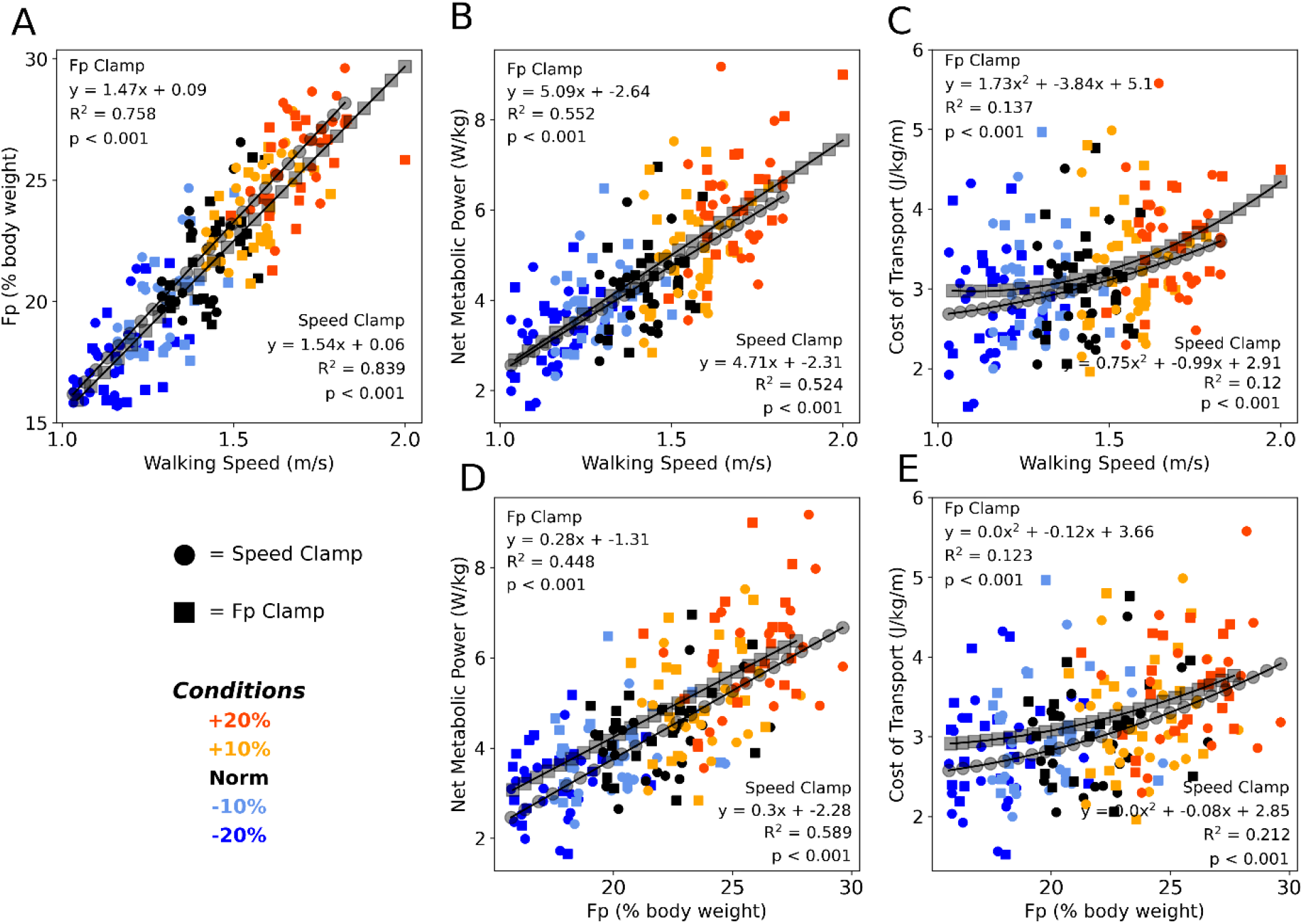
Correlations for both speed clamp (circles) and F_P_ clamp (squares) trials for each primary outcome variable. Walking speed correlated very strongly with F_P_ (A), moderately with net metabolic power (B), and very weakly with cost of transport (C). F_P_ correlated moderately with net metabolic power (D) and weakly with cost of transport (E). The polynomial and correlation coefficients were qualitatively similar between clamp types (circles & squares within each subplot) and across predictor variables (subplots B&D, C&E).

### Effects of Clamp Type

We found significant main effects of clamp type, where, on average across all condition intensities, the F_P_ clamp elicited 2.6% faster speeds, 1.4% greater F_P_, 8.9% higher net metabolic power, and 6.2% greater CoT compared to the speed clamp (Fig. 5, p≤0.01, η_p_^2^≥0.298). We also found significant interactions between condition intensity and clamp type for walking speed and F_P_. The interactions revealed that the difference between clamp types became larger with slower speed and with larger F_P_ (Fig. 5A-B). At the lowest condition intensity, participants walked faster (0.07±0.03 m/s *(mean difference ± standard deviation)*, p=0.008, d=0.887) and with higher net metabolic power (0.49±0.24 W/kg, p=0.044, d=0.660) during F_P_ clamp trails compared to speed clamp trials, despite exerting an indistinguishable F_P_ magnitude.

**Figure 5:**
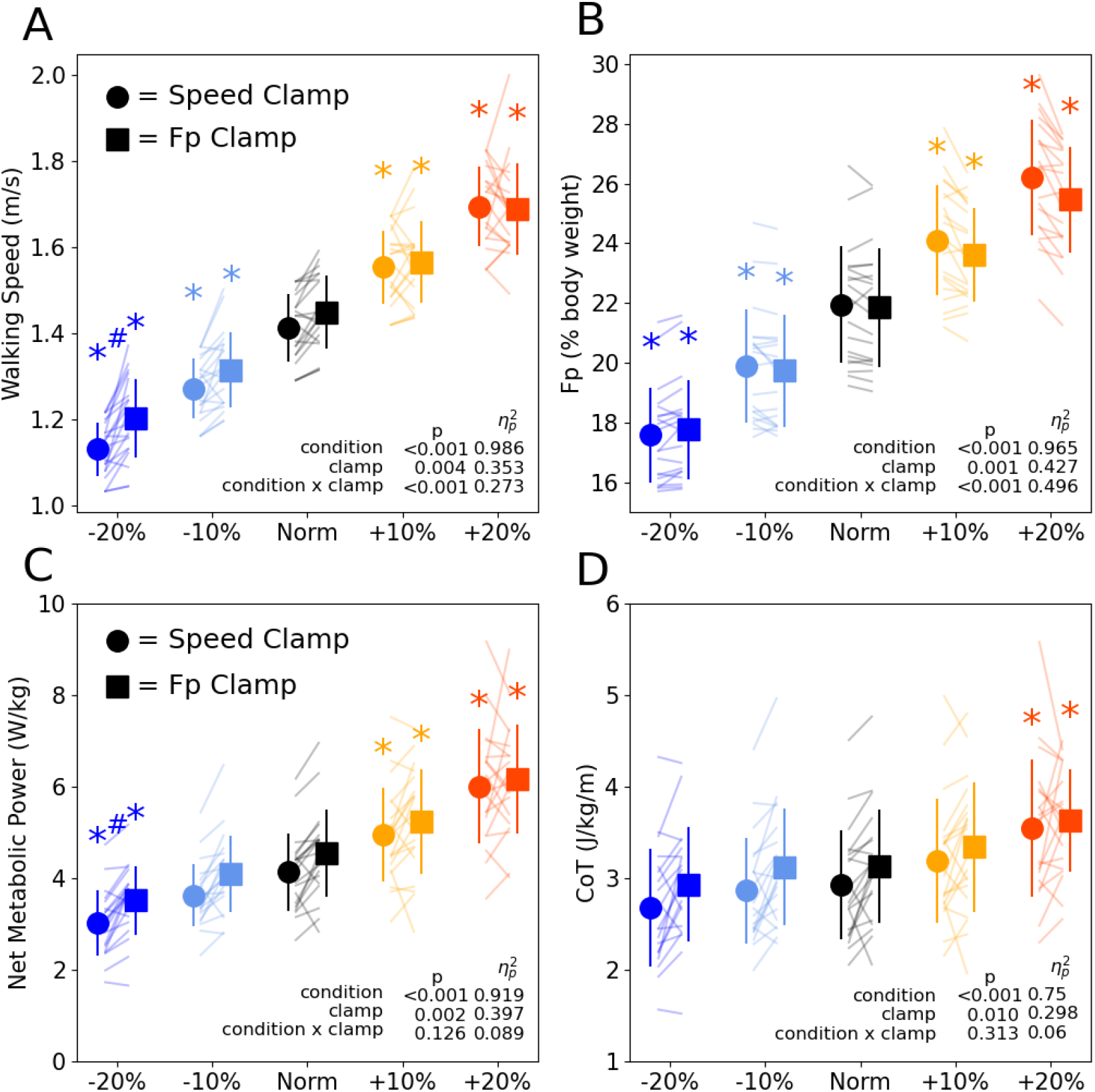
Walking speed (A) and F_P_ (B) across condition intensities for speed clamp and F_P_ clamp trials. Walking speed and F_P_ similarly increased and decreased with condition intensity. However, at the lowest condition intensity (−20%), particpants walked at faster speeds while extering the same F_P_. Walking net metabolic power (C) changed with condition intensity except for the -10% condition intensity. Walking cost of transport (D) only increased for the +20% condition intensity. Asterisks (*) indicate a significant pairwise post-hoc difference (p<0.05) from the Norm condition. Hashtags (#) indicate a significant pairwise post-hoc difference (p<0.05) between clamp types (speed vs F_P_).

### Effects of Condition Intensity

We found significant main effects of condition intensity for all primary outcome variables (F_P_, walking speed, net metabolic power, and CoT, Fig. 5). Participants increased and decreased their F_P_ and walking speed in response to higher and lower condition intensities, with each level significantly different from Norm across both clamp types (p≤0.002, d≥0.957, Fig. 5A-B). Net metabolic power increased and decreased along with changes in intensity for all conditions (p≤0.049, d≥0.642) except when prescribing the -10% condition intensity for both clamp types (p≥0.050, d≤0.639, Fig. 5C). Conversely, CoT only significantly increased from Norm when prescribing +20% condition intensity for both speed and F_P_ clamps (Fig. 5D, p≤0.013, d≥0.827).

## Discussion

Our goal was to objectively establish cause-effect relations between F_P_, walking speed, and walking metabolism across a range of intensities through a unique experimental paradigm designed to separately prescribe both F_P_ and walking speed in a cohort of healthy young adults. This protocol clamped (i.e., held steady) either F_P_ or walking speed, while measuring naturally emergent changes in the other as well as any effects on walking metabolism. Our group and others have observed indirect evidence alluding to cause-and-effect associations between F_P_ and walking speed and have reported metabolic consequences when deviating from preferred speed or F_P_ during typical walking.^13,16,17,22–27^ However, the recent proliferation of self-pace treadmill controllers^17,28,29^ provides a way to build upon these earlier observational studies. Consistent with the intuition established by prior work, we found a strong correlation between walking speed and F_P_, with ∼80% of variance explained between these factors. In addition to those correlations, we add empirical evidence showing that walking with a smaller or larger F_P_ demonstrably leads to the instinctive selection of slower or faster walking speeds, respectively.

This evidence in support of cause-effect relations provides valuable validation for individuals seeking to design and implement strategies to improve walking speed among older adults or individuals with gait pathology. Older adults and people with gait pathology walk at slower preferred speeds and with diminished push-off forces. Although reduced F_P_ has been implicated as a potential cause of slower walking speeds, their simultaneous presentation makes mechanistic insight difficult. With our documentation of the strong relation between F_P_ and walking speed, we can legitimately point to insufficient F_P_ generation as a root origin of slower walking speeds. We suspect that this relation would persist irrespective of the specific mechanism(s) giving rise to a diminished push-off in walking. Our results build confidence that interventions designed to augment F_P_ can be used to increase walking speed, as demonstrated by our prior research^30,31^ and relevant work from others^13,17,32,33^.

### Biomechanics in the Context of Walking Metabolism

Our secondary goal was to establish the metabolic consequences of the interplay between F_P_ and walking speed. We found two key outcomes regarding walking metabolism. First, we found that net metabolic power was more strongly associated with changes in walking speed and F_P_ (R^2^≈0.53, Fig. 4 B&D) compared to CoT (R^2^≈0.15, Fig. 4 C&E), at least across a prescribed change of ±20% in condition intensity. Previously, our group found that when walking at fixed speeds, both net metabolic power and CoT (inferred from equivalent speeds) increased by ∼20% when targeting 20% larger F_P_ and increased by ∼30% when targeting 20% smaller F_P_.^34^ Conversely, in this study, net metabolic power increased by ∼40% and decreased by ∼25% on average when targeting 20% larger and smaller F_P_, respectively. Additionally, when participants in this study could instinctively select their own walking speed, CoT increased by ∼16% on average when targeting 20% larger F_P_ but did not differ from usual walking when targeting 20% smaller F_P_. This relative lack of sensitivity of CoT to changes in condition intensity agrees with the “broad minimum” theory, wherein a range of walking speeds neighboring the local minimum of the CoT curve may share similar metabolic costs.^35^ For normal- and lower-intensity conditions, our participants adjusted F_P_ or speed in a manner that maintained a relatively invariant CoT, and thereby operated within their “broad minimum” CoT. Thus, under certain circumstances, walkers may exploit the interaction between F_P_ and walking speed to preserve or even reduce walking CoT. Ultimately, changing F_P_ or walking speed predictably alters net metabolic power but need not impact CoT.

Second, in further support of the strong relation between F_P_ and walking speed, we discovered that changing the magnitude of either yielded relatively similar effects on measures of walking metabolism. In other words, whether we prescribed a change in F_P_ or walking speed, effects on walking metabolic cost were nearly indistinguishable. We noted this also from our correlations (Fig. 4), which revealed quantitatively similar R^2^ values and regression coefficients as well as qualitatively similar trendlines between clamp types. However, this is not to say that there were no meaningful differences between clamp types. Indeed, we found a significant main effect of clamp type on metabolic cost; F_P_-clamp trials tended to require 9% higher net metabolic power and 6% CoT compared to speed-clamp trials on average (Fig. 5C-D). We have previously shown that walking with F_P_ biofeedback at a fixed treadmill speed does not itself exact a metabolic penalty.^34^ Nevertheless, we suspect that the greater metabolic energy cost associated with F_P_ clamp trials may arise from a cognitive “tax” levied to simultaneously regulate step-to-step adjustments in response to biofeedback and to treadmill self-pacing. Such a tax may allude to additional cognitive processing and/or neuromuscular costs associated with a shift toward supraspinal control of walking patterns rather than primarily relying on central pattern generators in the spinal cord. In this scenario, adjusting one’s F_P_ and walking speed in response to targeted biofeedback appears to require additional energy than simply walking at a fixed speed without engaging with biofeedback. Alternatively, the greater metabolic cost exhibited during the self-paced F_P_ clamp conditions may arise from periodic acceleration and deceleration of walking speed and subsequent metabolic effects.

### A Potential Discrepancy in the Speed-F_P_-Economy Relation

Because F_P_ and walking speed are inextricably linked, we would expect our protocol to yield highly similar biomechanical and metabolic outcomes across both clamp types. Indeed, this was true for most outcomes across most condition intensities (Fig. 5). However, when targeting 20% smaller than normal F_P_, our participants could have selected a slower walking speed and lower net metabolic power when producing the requisite F_P_, but they chose not to. Rather, we identified a naturally-emergent discrepancy between clamp types, in which participants exerted similar F_P_, but selected faster speeds at a higher net metabolic power during the F_P_ clamp than the speed-clamp. The instinctive selection of faster speeds at a metabolic penalty despite an indistinguishable F_P_ across clamp types demonstrates that humans do not *always* seek to minimize metabolic cost. In our daily lives, we may prioritize factors other than walking economy when we rush, become excited, or feel threatened or scared. However, we can see evidence of this even in laboratory environments. For example, healthy young adults sometimes select walking speeds somewhat faster than their most economical speed, even though they spend more energy when doing so.^35^ In another example, young healthy subjects have been shown to prioritize stability rather than take advantage of gravity-aided propulsion when walking down a gentle slope.^36^ These phenomena may explain the discrepancy we identified at low condition intensities, potentially optimizing a cost function other than the most economical gait patterns. We plan to further investigate how participants regulated their speed on a step-to-step basis when interacting with the self-pace controller, and the role that lower extremity muscles serve in generating and regulating F_P_.

### Translational implications

It is unclear whether these direct relations between F_P_, walking speed, and walking metabolism will hold in populations who may be candidates for clinical countermeasures to enhance gait performance or mitigate walking-related fatigue. For example, older adults typically walk slower, with smaller F_P_, and at higher metabolic costs compared to young adults. It is actually not well known whether or not older adults have movement biomechanics and underlying muscle actions that are tuned to minimize metabolic cost at their preferred speeds. Older adults also have the capacity to generate propulsive forces comparable to those measured in younger adults, but typically choose not to utilize that additional force capacity to increase walking speed.^23^ Future studies may consider enrolling older adults to participate in a similar design to determine whether age influences the cause-effect relation between F_P_ and walking speed, or if the metabolic consequences of that relation are altered by hallmark changes in muscle morphology and composition, cardiopulmonary function, sensorimotor integration, or executive processing.

### Limitations

One limitation in this study is that our F_P_ magnitudes and walking speeds were limited to a relatively small range compared to other studies that quantify walking metabolism. Our span across condition intensities deviated 20% from typical gait, yielding walking speeds between 1 and 2 m/s. Our protocol was informed by the magnitude of changes we would deem clinically meaningful. However, speed-dependent increases in CoT in otherwise healthy young adults do not typically arise until ≤1 m/s^9,10^. Another limitation is that we analyzed average profiles over the final 2 minutes of each 5-minute walking trial. Although subjects were provided an exploration period and reported being comfortable with biofeedback and self-pacing, further practice could have affected walking metabolic cost. Finally, we did not quantify individual determinants of propulsive force (i.e., trailing limb angle and peak ankle moment^18,22^).

## Conclusions

Using a unique clamp protocol, we confirm a direct causal relation between F_P_ and walking speed. We provide strong empirical evidence that young adults walking with larger/smaller F_P_ yields faster/slower walking speeds, demonstrating that the peak anterior component of the ground reaction force during push-off (F_P_) is an independent factor that governs walking speed. We also quantify the metabolic implications of altering walking speed and F_P_, finding that comparable changes in either F_P_ or walking speed elicit predictable and relatively uniform changes in walking metabolic cost.

## Data Availability

All data produced in the present study are available upon reasonable request to the authors

## Acknowledgments

Thank you to the people who volunteered to participate in this study. National Institutes of Health grant R01AG058615 provided funding for this study.

## Conflict of Interest Statement

The authors report that they have no conflicts to disclose.

## Notes

### Competing Interest Statement

The authors have declared no competing interest.

### Funding Statement

This study was funded by National Institutes of Health grant R01AG058615

### Author Declarations

The IRB of University of North Carolina at Chapel Hill gave ethical approval for this work.

